# Focal Usual Interstitial Pneumonia-like Fibrosis is a Core Prognostic Factor in Progressive Pulmonary Fibrosis

**DOI:** 10.1101/2023.12.07.23298650

**Authors:** Yukio Tsushima, Ethan N. Okoshi, Sousuke Ishijima, Andrey Bychkov, Kris Lami, Shimpei Morimoto, Yasuhiko Yamano, Kensuke Kataoka, Takeshi Johkoh, Yasuhiro Kondoh, Junya Fukuoka

**Author notes:** Address for correspondence: Junya Fukuoka MD, PhD, Department of Pathology, Nagasaki University Graduate School of Biomedical Sciences, 1-12-4 Sakamoto, Nagasaki 852-8523, Japan. Co-first authors (Yukio Tsushima, Ethan N. Okoshi).

## Abstract

Progressive pulmonary fibrosis (PPF) is a newly recognized clinical phenotype of interstitial lung diseases in the 2022 interstitial pulmonary fibrosis (IPF) guidelines. This category is based entirely on clinical and radiological factors, and the background histopathology is unknown. Our objective was to investigate the histopathological characteristics of PPF and to examine the correlation between usual interstitial pneumonia (UIP) and prognosis in this new disease type. We hypothesized that the presence of UIP like fibrosis predicts patients’ survival in PPF cases.

We selected 201 cases fulfilling the clinical criteria of PPF from case archives. Cases diagnosed as IPF by a multidisciplinary team were excluded. Whole slide images were evaluated by three pathologists who were blind to clinical and radiological data. We measured areas of UIP-like fibrosis and calculated what percentage of the total lesion area they occupied.

The presence of focal UIP-like fibrosis amounting to 10% or more of the lesion area was seen in 148 (73.6%), 168 (83.6%), and 165 (82.1%) cases for each pathologist respectively. The agreement of the recognition of UIP-like fibrosis in PPF cases was above κ = 0.6 between all pairs. Survival analysis showed that the presence of focal UIP-like fibrosis correlated with worsened survival under all parameters tested (p < 0.001).

The presence of UIP-like fibrosis is a core pathological feature of clinical PPF and its presence within diseased areas is associated with poorer prognosis. This study highlights the importance of considering the presence of focal UIP like fibrosis in the evaluation and management of PPF.

## INTRODUCTION

Progressive pulmonary fibrosis (PPF) is a newly defined clinical phenotype of fibrotic interstitial lung disease (ILD) introduced in Idiopathic Pulmonary Fibrosis and Progressive Pulmonary Fibrosis in Adults: An Official ATS/ERS/JRS/ALAT Clinical Practice Guideline (hereafter referred to as the 2022 guidelines) [1]. It serves as an update to the former clinical phenotype of progressive fibrosing interstitial lung disease (PF-ILD). PPF is defined as at least two of the following three criteria occurring within the past year with no alternative explanation in a patient with an ILD other than idiopathic pulmonary fibrosis (IPF): worsening symptoms, radiological progression, and physiological progression [1]. Whereas prior clinical practice guidelines recommended a period of evaluation of 2 years for PF-ILD, PPF can be identified within a follow-up period of only 1 year, which has benefits in prognostication and treatment [1,2].

The disease progression of ILDs manifesting PPF is similar to that of IPF, a chronic, fibrosing interstitial pneumonia of unknown cause [1]. One could consider PPF to be the terminology for other ILDs which closely follow the disease progression and clinical trajectory of IPF [3,4], which is reflected primarily by a decline in FVC, worsening of dyspnea, reduction in exercise capacity, and deterioration in health-related quality of life [5]. IPF is associated mainly with the radiological and histopathological features of usual interstitial pneumonia (UIP) [6], but UIP can also be present in ILDs other than IPF [7-9]. According to the established diagnostic criteria, UIP is histopathologically defined as an area of *1)* marked fibrosis/ architectural distortion, ± honeycombing in a predominantly subpleural/ paraseptal distribution; *2)* patchy involvement of lung parenchyma by fibrosis; *3)* fibroblast foci; and *4)* the absence of features that suggest an alternative diagnosis [10]. When only some of these findings are present but the morphology is still suggestive of UIP, a diagnosis of “probable” or “possible UIP” is appropriate [10,11].

The prevalence and progression of UIP associated features on computerized tomography (CT) and histology have been shown to predict mortality in patients with IPF [12-15]. Therefore, it is important to understand the histological features of PPF as well. However, the 2022 guidelines include no information regarding PPF’s histopathological presentation.

Previous studies have reported the coincidence of UIP and ILDs [7]. Specifically, Flaherty et al. studied the coincidence of UIP with non-specific interstitial pneumonia (NSIP), terming this condition “discordant UIP”. Discordant UIP showed slightly better survival than concordant UIP but this difference in survival did not reach statistical significance [7]. Thus, subsequent guidelines avoided subcategorizing discordant UIP within the wider category of UIP. In the present study, we intended to investigate further the phenomenon of discordant UIP within the context of PPF’ hypothesizing that even if the area of UIP was not large enough to merit a diagnosis of UIP under the standard criteria, the presence of trace discordant UIP would still have some effect on prognosis in PPF cases.

The aim of this study was firstly to investigate the histopathological characteristics of PPF with a focus on determining the prevalence of UIP pattern or focal UIP-like fibrosis in the pathology of PPF, and secondly to examine the correlation between focal UIP-like fibrosis and prognosis in relation to underlying etiology. Given the clinical similarities between IPF and PPF, we hypothesized that UIP may constitute a fundamental histological aspect of PPF.

## MATERIALS AND METHODS

### PATIENT DATA

A total of 201 cases fulfilling the clinical criteria of PPF as evidenced by pulmonary function tests, patient symptoms, or radiological images from 2009 - 2018 were enrolled from a case archive at Tosei General Hospital (Aichi, Japan) and confirmed via multidisciplinary discussion (MDD). Specifically, pulmonary function tests included measuring FVC and DLco over time [1]. Since the 2022 guidelines describe PPF as a distinct category of ILDs separate from IPF, we excluded all cases diagnosed as IPF via MDD from our cohort (Figure 1). Clinical data and whole slide images of tissue sections derived from surgical lung biopsies (SLB) were collected for each patient (Aperio CS2, 20x magnification, Leica Biosystems, Germany). Typically, one biopsy each was taken from segments S5, S8, and S9, and the average number of slides per case was 2.65. There were 146 patients with all 3 lobes sampled, 21 patients with only S5 and S8, 3 patients with only S5 and S9, 16 patients with only S8 and S9, and for 5 patients each which only had 1 of the 3 lobes sampled. This study was carried out at a single institution in compliance with the principles of the Declaration of Helsinki and approved by its institutional review board (IRB No. 14012746, February 3, 2014).

**Figure 1.**
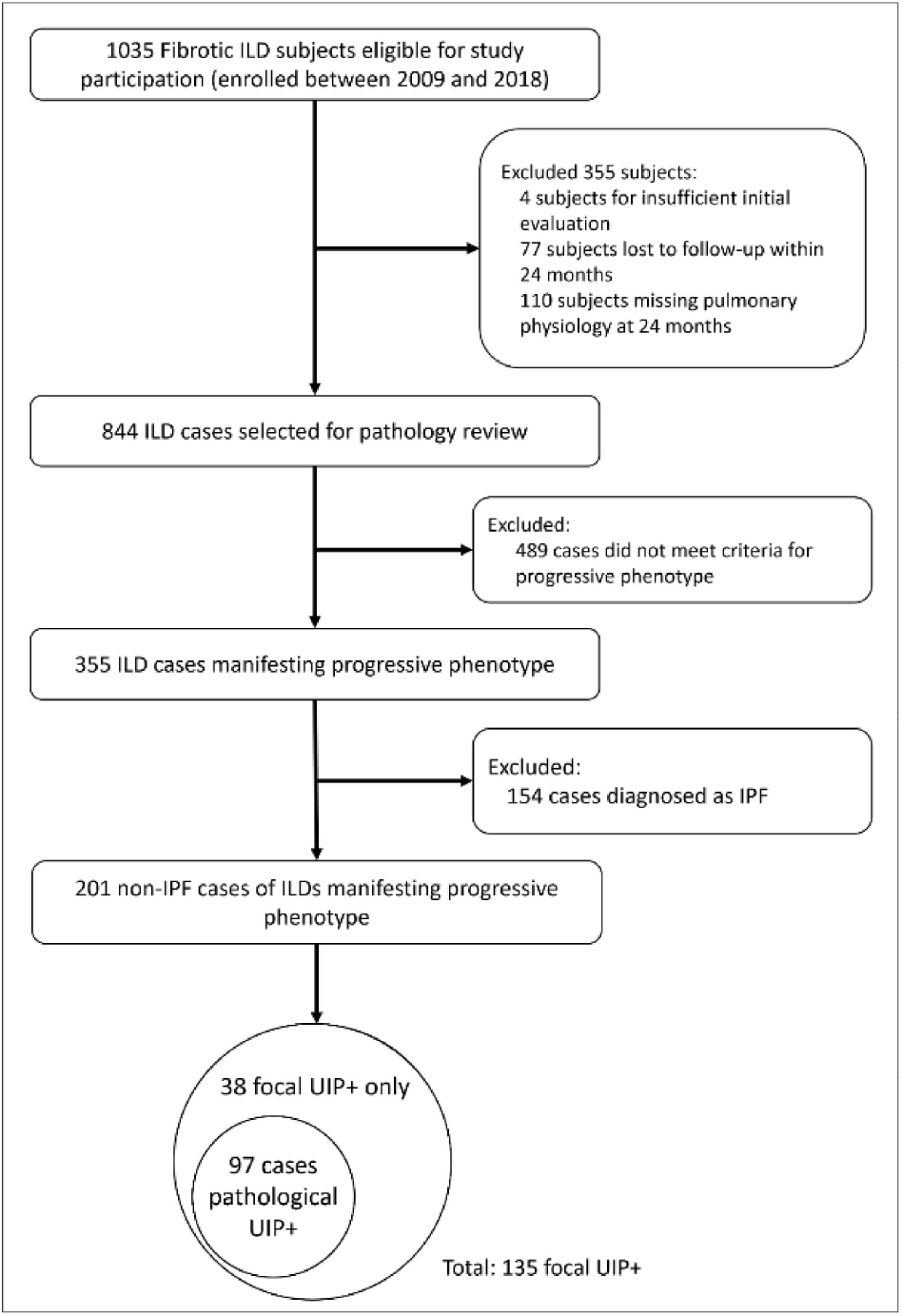
Pipeline Diagram Describing Study Cohort. 201 cases of non-IPF ILDs exhibiting progressive phenotype were collected from Tosei General Hospital between 2009 and 2018. These cases were first labeled as pathological UIP+ or pathological UIP−. We then expanded the criteria for UIP positivity, labeling any UIP which was over 10% of the total disease area as “focal UIP”. Using the criteria for focal UIP, we labeled an additional 38 cases as focal UIP+ for a total of 135 cases positive for focal UIP. IPF, idiopathic pulmonary fibrosis; ILD, interstitial lung disease; UIP, usual interstitial pneumonia.

### DIGITAL HISTOLOGICAL ANALYSIS

WSIs were evaluated by three pathologists (YT, JF, SI) independently, blinded to both clinical and radiological data. The areas of UIP like fibrosis were demarcated using a digital pen tool (Aperio ImageScope, Leica Biosystems, Germany). The area of UIP-like fibrosis and the total area of ILD were measured, excluding normal tissue area in all specimens obtained through SLB. The focal area of UIP-like fibrosis was defined as the area of UIP divided by the total lesion area (**Supplementary Figure 1**).

Two different labels were used for marking cases as positive or negative for UIP, the first being “pathological UIP”, or the histological criteria outlined in the 2010 IPF guidelines for definite, probable, or possible UIP, and the second being “focal UIP”, which we defined as UIP pattern which was greater than or equal to 10% ofthe total area ofILD (**Figure 2**).

**Figure 2.**
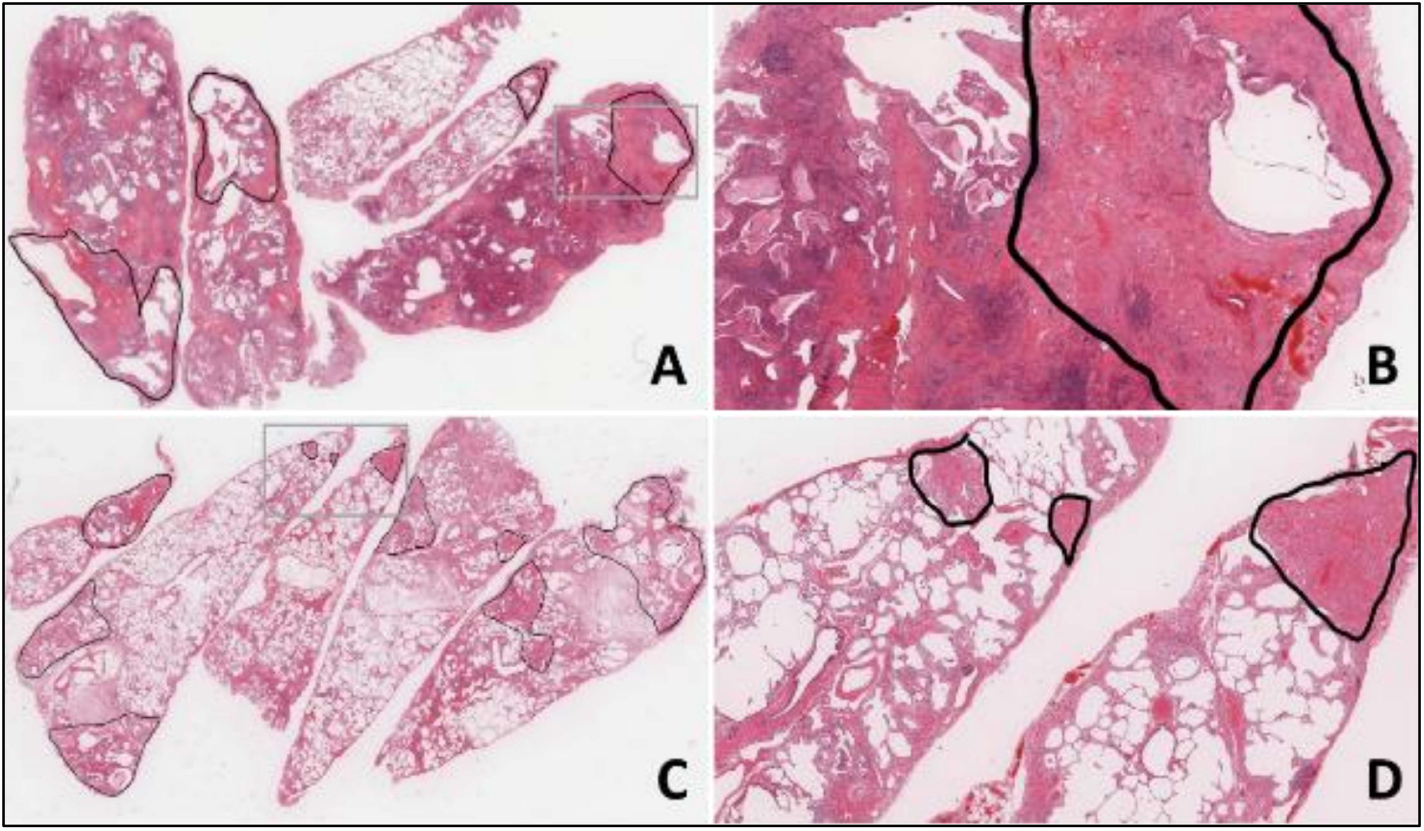
Representative HE images of areas of UIP. UIP was defined as destructive fibrosis often with honeycombing that did not reach the threshold ofNSIP and did not necessarily include fibroblastic foci or temporal heterogeneity, as per the 2010 IPF guidelines. The case depicted in A and B shows UIP occupying 28.3% of the diseased area, while the case depicted in C and D shows UIP occupying 17.0% of the diseased area. Annotated areas demarcated with a black line indicate areas of UIP. The gray box indicates areas that were enlarged on the right. HE, hematoxylin and eosin stain; UIP, usual interstitial pneumonia.

To elaborate, focal UIP was defined as destructive fibrosis that did not reach the threshold of NSIP, and did not necessarily include fibroblastic foci or temporal heterogeneity [1,6,10,11]. Due to the focal nature of this category, there is a high likelihood that patchiness or fibroblastic foci are missing, thus many ofour focal UIP+ cohort (UIP-like fibrosis ≥ 10% of disease area) would fall under the diagnosis of “probable” or “possible UIP”, according to the 2010 IPF guidelines. We decided to use the 2010 IPF guidelines over more recent editions because the language concerning UIP positivity was clearer than that of the 2018 or 2022 guidelines, as these later guidelines include the category of “indeterminate for UIP”, which includes consideration for features favoring UIP secondary to another cause [1,10,11].

The cutoff value for focal UIP was determined using receiver operating characteristic (ROC) analysis and interobserver agreement was evaluated using Cohen’s kappa. Specifically, we evaluated cutoff values ranging from 10% to 50% area of focal UIP-like fibrosis, and a threshold of 10% showed both the best agreement between the 3 pathologists and separation of prognostic difference between UIP and non-UIP (**Supplementary Figure 2**). For cases with multiple biopsy specimens, we took the average percentage across specimens. In cases where the three pathologists differed on the diagnosis of UIP content, the final diagnosis was made by consensus.

### STATISTICAL ANALYSIS

Categorical data is presented as counts and frequencies (percentages). Continuous variables are presented as median ± IQR. Statistical comparisons between categorical groups were performed using Pearson’s Chi-squared test for variables with all expected values ≥5 and Fisher’s exact test for expected values <5. Comparisons between categorical and continuous data were conducted using the Wilcoxon rank sum test. The patients’ survival data was analyzed through a log-rank test, and Kaplan-Meier curves were generated based on the scores of each pathologist. Overall survival from surgery to death (n = 50) or lung transplant (n = 5) was analyzed by univariate and multivariate Cox proportional hazards regression analyses. Results are expressed as hazard ratios (HRs), their 95% confidence intervals, and associated Wald test p values. An association between a time-to-event variable and a candidate for a predictor of the event was shown as a p-value in the log-rank test, the hazard ratio, and the Kaplan-Meier curves. The inconsistencies between disease groups in the association between focal UIP positivity and hazard ratio were assessed using the empirical distribution of the hazard ratio obtained from data with randomized disease labels. Empirical distributions of the hazard ratio for each disease group were constructed from 2000 random permutations of the disease labels which were then used to fit a Cox proportional hazards model. All analysis was conducted in R (4.3.0).

## RESULTS

### PATIENT POPULATION

Patients’ baseline demographic, clinical, and physiological characteristics are shown in **Table 1**. Analysis showed that men were statistically more likely to be identified as pathological UIP+. Age was a statistically significant predictor for pathological UIP, with positive patients tending to be older than pathological UIP-patients. Smoking history was seen to significantly differ between the focal UIP groups but did not significantly differ in the pathological UIP groups. The distribution of smokers vs. nonsmokers by sex was as follows: for men, 76 current or ex smokers and 16 nonsmokers, and for women, 24 current or ex-smokers and 85 nonsmokers. Unexpectedly, in our cohort, focal UIP+ patients exhibited significantly higher FVC values than those without, whereas pathological UIP+ patients exhibited significantly higher FVC% values than those pathological UIP-. We found no significant differences in DLco, DLco%, or KL-6 expression between pathological UIP+/-groups or focal UIP+/-groups. There was no significant correlation between the number of sampled lobes and focal or pathological UIP positivity (p = 0.6).

**Table 1.** Baseline Characteristics in 201 Patients with Progressive Pulmonary Fibrosis Who Underwent Surgical Lung Biopsy. Significant p-values are highlighted with bold text. Significance was evaluated at p < 0.05. Categorical values presented as counts and row-wise frequency. Continuous variables presented as mean ± standard deviation. UIP, usual interstitial pneumonia; cHP, chronic hypersensitive pneumonia; CTD-ILD, connective tissue disease-associated interstitial lung disease; iNSIP, idiopathic non-specific interstitial pneumonia; iPPFE, idiopathic pleuroparenchymal fibroelastosis; UC-ILD, unclassifiable interstitial lung disease. Pathological UIP = definite, possible, or probable UIP according to the 2010 IPF guidelines. Focal UIP = UIP-like fibrosis comprises≥, 10% of disease area.

| Characteristic | Pathological UIP |  |  | Focal UIP |  |  | Total<br>N = 201<sup>1</sup> |
| --- | --- | --- | --- | --- | --- | --- | --- |
|  | Negative, N = 104<sup>1</sup> | Positive, N = 97<sup>1</sup> | p-value | Negative, N = 66<sup>1</sup> | Positive, N = 135<sup>1</sup> | p-value |  |
| Sex |  |  | 0.061<sup>2</sup> |  |  | <0.001<sup>2</sup> |  |
| F | 63 (58%) | 46 (42%) |  | 48 (44%) | 61 (56%) |  | 109 (100%) |
| M | 41 (45%) | 51 (55%) |  | 18 (20%) | 74 (80%) |  | 92 (100%) |
| Age | 61 (55, 66.25) | 63 (60.00, 68) | **0.021<sup>3</sup>** | 62 (55, 67) | 62 (58, 67) | 0.2<sup>3</sup> | 62 (57, 67) |
| Smoking history |  |  | 0.5<sup>2</sup> |  |  | **0.004<sup>4</sup>** |  |
| current | 6 (55%) | 5 (45%) |  | 3 (27%) | 8 (73%) |  | 11 (100%) |
| ex | 42 (47%) | 47 (53%) |  | 19 (21%) | 70 (79%) |  | 89 (100%) |
| never | 56 (55%) | 45 (45%) |  | 44 (44%) | 57 (56%) |  | 101 (100%) |
| FVC | 2.21 (1.72, 2.72) | 2.36 (2.01, 3.09) | 0.061<sup>3</sup> | 2.03 (1.67, 2.67) | 2.41 (1.95, 3.09) | **0.004<sup>3</sup>** | 2.30 (1.82, 2.92) |
| %FVC | 81 (69, 95) | 87 (75, 98) | **0.038<sup>3</sup>** | 79 (69, 93) | 86 (72, 99) | 0.052<sup>3</sup> | 85 (71, 97) |
| DLco | 10.2 (8.5, 13.5) | 10.6 (8.7, 12.5) | 0.7<sup>3</sup> | 9.9 (8.8, 11.6) | 11.1 (8.5, 13.3) | 0.3<sup>3</sup> | 10.4 (8.5, 13.0) |
| Missing | 2 | 0 |  | 2 | 0 |  | 2 |
| %DLco | 61 (49, 75) | 65 (51, 77) | 0.6<sup>3</sup> | 62 (49, 74) | 64 (50, 78) | 0.7<sup>3</sup> | 63 (49, 76) |
| Missing | 2 | 0 |  | 2 | 0 |  | 2 |
| KL-6 | 1,431 (816, 2,453) | 1,102 (663, 2,159) | 0.2<sup>3</sup> | 1,507 (861, 2,543) | 1,102 (662, 2,241) | 0.065<sup>3</sup> | 1,204 (726, 2,380) |
| Disease |  |  | **0.018<sup>4</sup>** |  |  | 0.6<sup>4</sup> |  |
| cHP | 5 (36%) | 9 (64%) |  | 4 (29%) | 10 (71%) |  | 14 (100%) |
| CTD-ILD | 47 (57%) | 35 (43%) |  | 30 (37%) | 52 (63%) |  | 82 (100%) |
| iNSIP | 15 (79%) | 4 (21%) |  | 8 (42%) | 11 (58%) |  | 19 (100%) |
| iPPFE | 1 (50%) | 1 (50%) |  | 0 (0%) | 2 (100%) |  | 2 (100%) |
| UC-ILD | 36 (43%) | 48 (57%) |  | 24 (29%) | 60 (71%) |  | 84 (100%) |

<sup>1</sup> n (%); Median (IQR)<sup>2</sup> Pearson's Chi-squared test<sup>3</sup> Wilcoxon rank sum test<sup>4</sup> Fisher's exact test

We then performed a logistic analysis on the patient cohort statistics to test for correlation with survival. In the consensus analysis, no factors were found to correlate with improved or worsened survival (**Supplementary Table 1**).

### HISTOLOGY

Primary histological findings upon analysis of our cohort are described in **Table 2**. The most frequently identified histological pattern was NSIP, with 38.8% of our cohort exhibiting this pattern. Following that, definite and probable UIP were the next most common findings with 31 cases and 33 cases respectively (15.4% and 16.4%). There were 13 total separate findings identified in the histology of our cohort, with only 4 of these findings present in more than 10% of the cohort.

**Table 2.** Histopathological findings in a 201 Patient PPF Cohort. Only primary findings are reported. NSIP, non-specific interstitial pneumonia; UIP, usual interstitial patteren; ALI, acute lung injury; DAD, diffuse alveolar damage; ACIF, airway-centered interstitial fibrosis; OP, organizing pneumonia; NOS, not otherwise specified; PPFE, pleuroparenchymal fibroelastosis; DIP, desquamative interstitial pneumonia; LIP, lymphocytic interstitial pneumonia; PAP, pulmonary alveolar proteinosis. Values shown as counts and frequency within cohort.

| Histology | Count | Frequency |
| --- | --- | --- |
| NSIP | 77 | 38.8% |
| Definite UIP | 31 | 15.4% |
| Probable UIP | 33 | 16.4% |
| Possible UIP | 16 | 8.0% |
| ALI/DAD | 23 | 11.4% |
| ACIF | 8 | 4.0% |
| OP | 3 | 1.5% |
| PPFE | 2 | 1.0% |
| Small airway disease | 2 | 1.0% |
| DIP | 1 | 0.5% |
| LIP | 1 | 0.5% |
| PAP | 1 | 0.5% |
| IP, NOS | 2 | 1.0% |

### PATHOLOGICAL UIP COHORT

Of the 201 total cases, 97 (48.3%) displayed a histopathological pattern consistent with the 2010 IPF guideline [10] definitions of definite UIP, probable UIP, possible UIP, or UIP vs. other (Table 3). The mean percentage of UIP pattern within the lesion area was 49.21% among these pathological UIP+ cases. Among all PPF cases, those labeled pathological UIP+ had a poorer prognosis than those that were not marked positive (p = 0.0005) (Figure 3). Overall survival (OS) at 36 months was 95% for the pathological UIP-group and 86% for the pathological UIP+ group, and at 60 months OS decreased to 88% and 67% respectively. At 120 months, OS was 73% and 49% respectively.

**Table 3.** Diagnosis Information for Pathological UIP in a 201 Patient PPF Cohort. Percent UIP+ in this table refers only to pathological UIP cases, not focal UIP cases. iNSIP, idiopathic non-specific interstitial pneumonia; cHP, chronic hypersensitive pneumonia; CTD-ILD, connective tissue disease-associated interstitial lung disease; iPPFE, idiopathic pleuroparenchymal fibroelastosis; UC-ILD, unclassifiable interstitial lung disease; UIP, usual interstitial pneumonia; PPF, progressive pulmonary fibrosis.

| PPF; Progressive Pulmonary Fibrosis (n = 201) |  |  |  |  |  |  |
| --- | --- | --- | --- | --- | --- | --- |
| Etiology | N | Definite UIP | Probable UIP | Possible UIP | UIP vs. Other | Percent UIP+ |
| iNSIP | 19 | 0 | 0 | 0 | 4 | 21% |
| cHP | 14 | 5 | 1 | 2 | 1 | 64% |
| CTD-ILD | 82 | 8 | 16 | 3 | 8 | 43% |
| iPPFE | 2 | 0 | 0 | 0 | 1 | 50% |
| UC-ILD | 84 | 13 | 16 | 11 | 8 | 57% |

**Figure 3.**
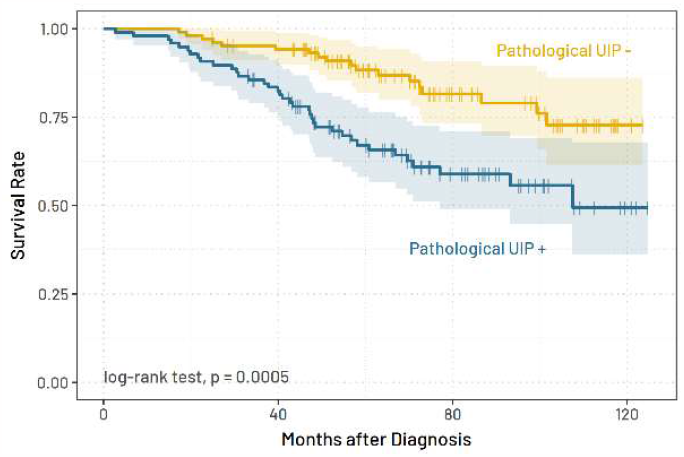
Kaplan-Meier survival curves for pathological UIP+ and pathological UIP− patients. Vertical ticks indicate last follow up visit. The yellow line indicates pathological UIP–cases (n = 104) and the blue line indicates pathological UIP+ cases (n = 97). UIP, usual interstitial pneumonia; PPF, progressive pulmonary fibrosis.

### FOCAL UIP COHORT

To evaluate different cutoff thresholds for focal UIP positivity, a Cox proportional hazards model was univariately fit to cutoff thresholds from 10% to 50%, and a 10% cutoff showed the greatest prognostic value (**Supplementary Figure 2A**). ROC analysis showed that the 10% cutoff threshold also had the best tradeoff between sensitivity and specificity among all cutoff thresholds ranging from 0% to 100% (**Supplementary Figure 2B**). Thus, the label of “focal UIP+” was applied to cases where the area of UIP comprised greater than 10% of the total disease area.

Among the 201 PPF cases, focal UIP was identified in 148 (73.6%), 165 (82.1%), and 168 (83.6%) cases by three experienced pathologists respectively. By consensus, 135 (67.1%) cases were established to be focal UIP+. The mean percentage of UIP area within a diseased area for a focal UIP+ case was 47.64%, and for cases that were pathological UIP-but then were labeled focal UIP+, the mean percentage of UIP area was 39.17%. Interobserver agreement was calculated to be 0.61, 0.63, and 0.71 (Cohen’s kappa) and deemed acceptable at the 10% cutoff value. There were 72 focal UIP+ cases which had a UIP area less than 50% of the disease area and 38 focal UIP+ cases which had a UIP area less than 30% of the disease area. Kaplan-Meier curves for each pathologist’s diagnoses of focal UIP+/-show that focal UIP+ cases have a significantly reduced survival compared to focal UIP–cases (Figure 4A). We also conducted survival analysis of the consensus determinations of focal UIP+/-, the result of which also showed decreased survival for focal UIP+ patients (**Figure 4B**). OS for consensus focal UIP− and focal UIP+ patients respectively was 97% and 87% at 36 months, 95% and 69% at 60 months, and 87% and 48% at 120 months.

**Figure 4.**
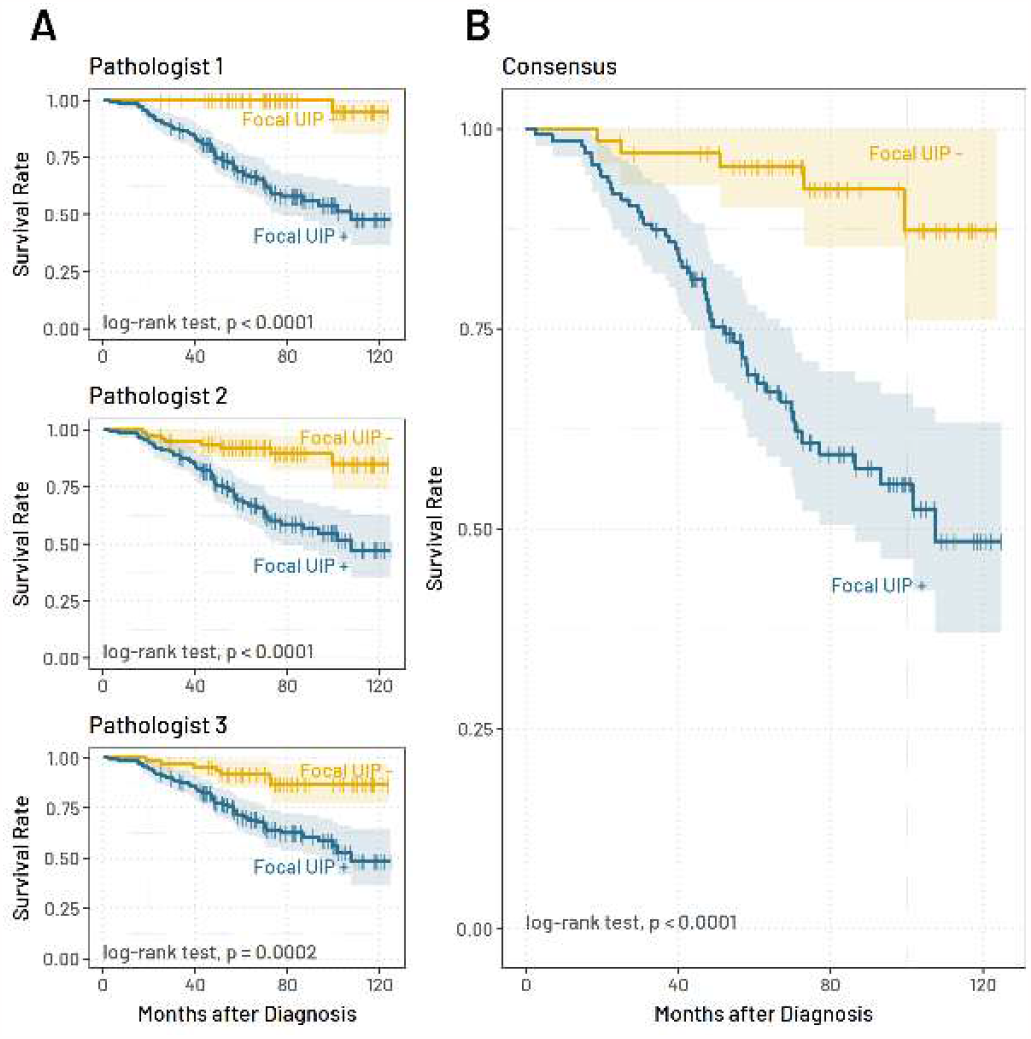
Kaplan-Meier survival curves for focal UIP+ and focal UIP–patients, separated by pathologist (A) and by consensus (B). Pathological UIP positivity or negativity was not a factor in this analysis – only focal UIP positivity or negativity was considered. Vertical ticks indicate last follow up visit. The yellow line indicates focal UIP–cases and the blue line indicates focal UIP+ cases. UIP, usual interstitial pneumonia.

### ETIOLOGY

Finally, we compared the survival of PPF with or without focal UIP in individual ILD etiologies. Out of 201 cases of PPF, the case counts were as follows: 19 idiopathic NSIP (iNSIP), 14 chronic hypersensitivity pneumonitis (cHP), 82 connective tissue disease-associated interstitial lung disease (CTD-ILD), 2 idiopathic pleuroparenchymal fibroelastosis (iPPFE), and 84 unclassifiable interstitial lung disease (UC ILD) (**Table 1**). Separated by etiology, a pathological UIP pattern was observed in 64% of cHP cases, 57% of UC-ILD cases, 43% of CTD ILD cases, 50% of iPPFE, and 21% of iNSIP cases (**Table 3**). Regardless of the underlying ILD etiology of PPF, patients with focal UIP consistently exhibit a poorer prognosis compared to those without, with statistical significance achieved in three out of four disease categories tested (CTD-ILD, NSIP, and UC-ILD) (**Figure 5**). Survival analysis for iPPFE was excluded due to the low case count (n=2). Interestingly, even in cases that were pathological UIP-, the presence of focal UIP fibrosis exceeding 10% tends to result in a worse prognosis (p = 0.0018) (**Figure 6**).

**Figure 5.**
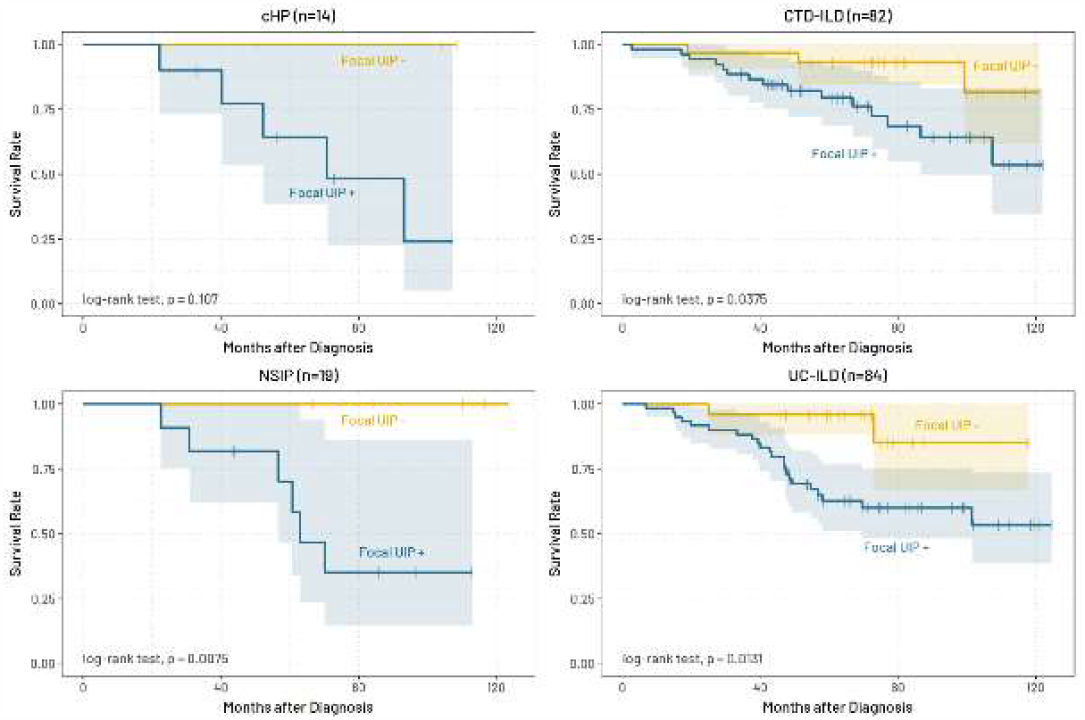
Kaplan Meier survival curves for focal UIP+ and focal UIP− patients, separated by etiology. Vertical ticks indicate last follow up visit. The yellow line indicates focal UIP–cases and the blue line indicates focal UIP+ cases. UIP, usual interstitial pneumonia.

**Figure 6.**
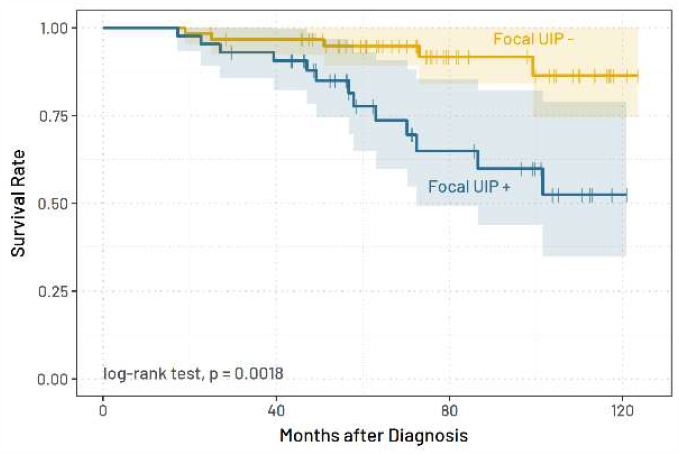
Kaplan-Meier survival curves for focal UIP+ and negative patients within pathological UIP–cases (n= 104). Vertical ticks indicate last follow up visit. The yellow line indicates focal UIP–cases (n=66) and the blue line indicates focal UIP+ cases (n=38). UIP, usual interstitial pneumonia.

### COX PROPORTIONAL HAZARDS MODEL

We performed univariate and multivariate Cox proportional hazards regression analysis to examine the effect of each clinical variable on patient survival. The multivariate Cox proportional hazards model showed that focal UIP positivity (HR = 6.43, p = 0.00015) and KL-6 value (HR = 0.56, p = 0.005) were the only significantly prognostic factors when controlling for focal UIP+/-, disease (i.e., cHP, CVD, NSIP, and PPFE), FVC, %FVC, DLco, %DLco, KL-6, smoking history, sex, and age. (**Table 4**). Harrel’s C-index for the model was 0.762 ± 0.036 and the global likelihood ratio test p-value was 0.000002. Univariate analysis of each of the above variables individually showed that focal UIP (HR = 6.24, p = 0.000096), sex (HR = 1.86, p = 0.025), DLco (HR = 0.92, p = 0.036), and %DLco (HR = 0.98, p = 0.037) were significantly prognostic.

**Table 4.** Cox proportional hazards regression analysis results. Factors were tested individually with multiple univariate Cox proportional hazards models and also together in a single multivariate Cox proportional hazards model. Two cases had missing DLco data. UIP, usual interstitial pneumonia; cHP, chronic hypersensitive pneumonia; CTD-ILD, connective tissue disease-associated interstitial lung disease; iNSIP, idiopathic non-specific interstitial pneumonia; iPPFE, idiopathic pleuroparenchymal fibroelastosis; UC-ILD, unclassifiable interstitial lung disease.

| Characteristic | N | Univariate |  |  | Multivariate |  |  |
| --- | --- | --- | --- | --- | --- | --- | --- |
|  |  | HR<sup>1</sup> | 95% CI<sup>1</sup> | p-value<sup>2</sup> | HR<sup>1</sup> | 95% CI<sup>1</sup> | p-value<sup>2</sup> |
| Focal UIP | 201 | 6.24 | [2.49, 15.7] | **<0.001** | 6.43 | [2.49, 16.6] | **<0.001** |
| Sex | 201 | 1.86 | [1.08, 3.19] | **0.025** | 2.94 | [0.68, 12.7] | 0.15 |
| Age | 201 | 1.02 | [0.99, 1.06] | 0.2 | 1.00 | [0.95, 1.06] | >0.9 |
| Smoking history | 201 |  |  |  |  |  |  |
| current |  | — | — |  | — | — |  |
| ex-smoker |  | 1.55 | [0.37, 6.49] | 0.6 | 3.22 | [0.71, 14.7] | 0.13 |
| never |  | 1.14 | [0.27, 4.81] | 0.9 | 4.05 | [0.78, 21.2] | 0.10 |
| FVC | 201 | 0.90 | [0.64, 1.26] | 0.5 | 0.73 | [0.12, 4.25] | 0.7 |
| %FVC | 201 | 0.99 | [0.97, 1.00] | 0.051 | 0.99 | [0.94, 1.05] | 0.8 |
| DLco | 199 | 0.92 | [0.84, 0.99] | **0.036** | 0.88 | [0.69, 1.13] | 0.3 |
| %DLco | 199 | 0.98 | [0.97, 1.00] | **0.037** | 0.99 | [0.95, 1.04] | 0.7 |
| KL-6 | 201 | 0.84 | [0.61, 1.15] | 0.3 | 0.56 | [0.37, 0.84] | **0.005** |
| Disease | 201 |  |  |  |  |  |  |
| cHP |  | — | — |  | — | — |  |
| CTD-ILD |  | 0.52 | [0.19, 1.41] | 0.2 | 0.56 | [0.18, 1.75] | 0.3 |
| iNSIP |  | 0.68 | [0.21, 2.23] | 0.5 | 0.58 | [0.15, 2.35] | 0.4 |
| iPPFE |  | 2.62 | [0.30, 22.7] | 0.4 | 5.13 | [0.45, 58.9] | 0.2 |
| UC-ILD |  | 0.79 | [0.30, 2.06] | 0.6 | 0.75 | [0.25, 2.31] | 0.6 |

<sup>1</sup> HR = Hazard Ratio, CI = Confidence Interval
<sup>2</sup> Wald test

### ASSOCIATION BETWEEN FOCAL UIP POSITIVITY AND THE SURVIVAL RATE

The heterogeneity between the disease groups, in the association between focal UIP positivity and the hazard ratio, were shown via referring histograms generated from the hazard ratios obtained from generating empirical distributions of the hazard ratios (**Figure 7**). The result showed that the estimated hazard ratios for focal UIP were significantly larger in the disease groups of cHP and iNSIP than in CTD-ILD and UC-ILD. The hazard ratio in the disease group of UC-ILD was the lowest among the disease groups. It should be noted that the distribution of p-values suggests high instability in the estimation of hazard ratios in the disease groups of cHP and iNSIP, likely due to low sample size.

**Figure 7.**
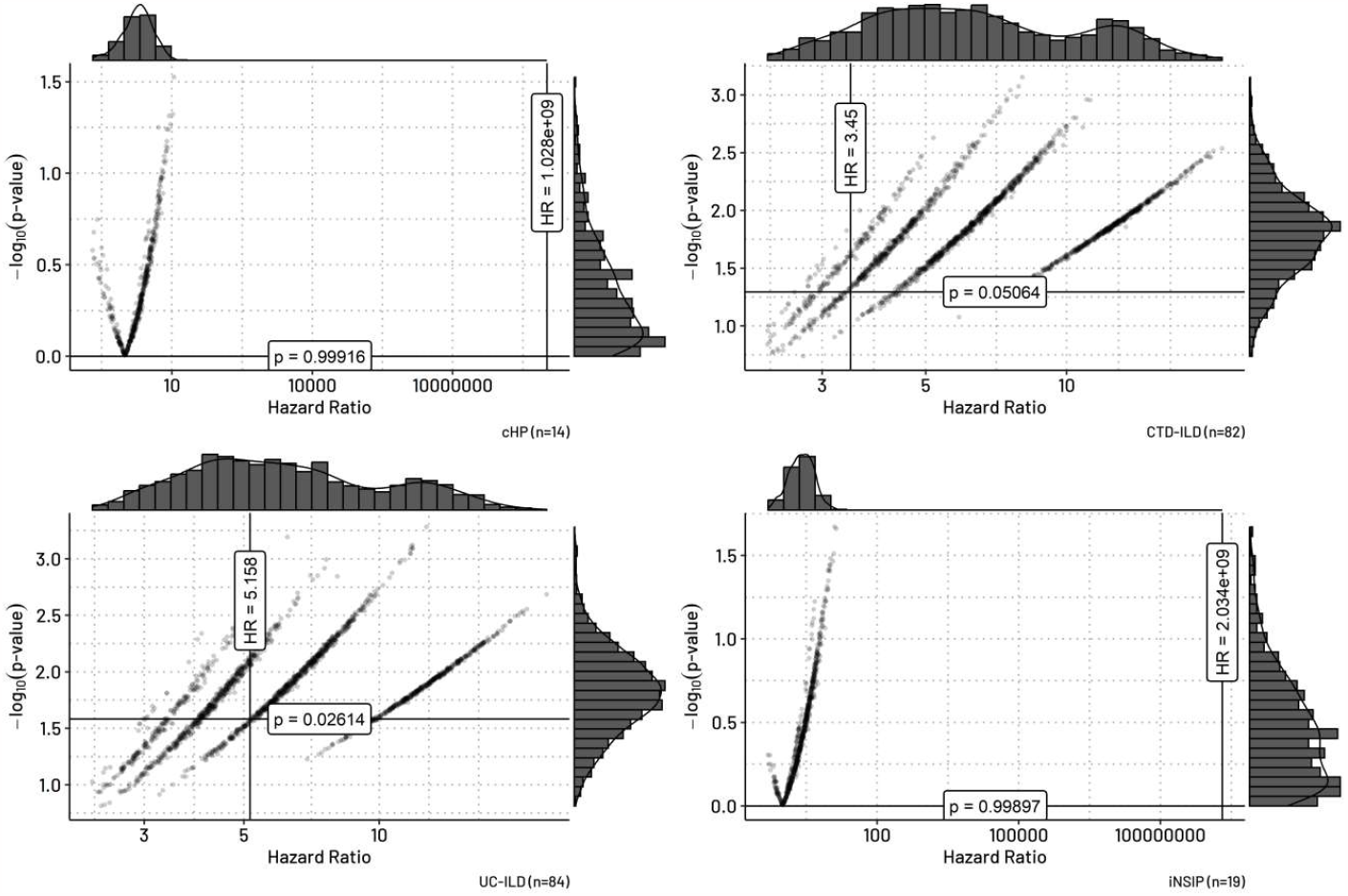
Permutation test results showing the empirical distribution of hazard ratios and p-values from 2000 random permutations of the disease type labels in a Cox proportional hazards model, separated by etiology. The vertical and horizonal lines in each plot show the hazard ratio and p-value of the cox proportional hazards model calculated from the real data. The histograms on the top and the on the right of each panel show the frequencies of the hazard ratios and the p values obtained from the results of 2000 permutations of the disease labels. iNSIP, idiopathic non-specific interstitial pneumonia; cHP, chronic hypersensitive pneumonia; CTD-ILD, connective tissue disease-associated ILD; iPPFE, idiopathic pleuroparenchymal fibroelastosis; UC-ILD, unclassifiable ILD.

## DISCUSSION

The 2022 IPF guidelines detailed the clinical and radiological criteria necessary to diagnose PPF in fibrotic ILDs but omitted histopathological criteria due to a lack of previously published data [1]. Identifying the histological features which correlate to disease progression is critical to deepening our understanding of this disorder. Interestingly, the physiological criteria for PPF were derived from IPF cases, but this extrapolation was not extended to include histopathological criteria. Thus, we hypothesized that UIP (i.e., the histological marker of IPF) would also hold significance in the histopathology of PPF. Our findings showed that the presence of UIP pattern in ILDs manifesting PPF correlated with worsened survival and had prognostic significance regardless of the underlying etiology.

Initially, we believed that UIP could be the dominant pathology of PPF, but this was not the case as it was identified in only half of our cohort. It was only when we expanded our criteria for UIP positivity, using the 10% cutoff value, that we found that two thirds of our PPF cases fell under the focal UIP+ label. While there are many other histologies included in PPF, we believe that focal UIP-like fibrosis could act as the core of PPF histology. For most cases in our cohort, it was the key to predicting prognosis, and in the future could guide treatment. However, one third of our PPF cohort were focal UIP-, and yet still exhibited progressive fibrotic disease enough to be classified as PPF.

Interobserver agreement was deemed to be overall encouraging as our Cohen’s kappa values were above 0.6 between all pairs, which is substantial agreement. Prior studies in interobserver variability and agreement in lung diseases shows that the diagnostic concordance of UIP is expected at most to be 0.5, and that values above this should be regarded positively [16-18].

The statistical significance in the difference between male and female rates of UIP positivity was understandable due to likewise significantly different rates in smoking history between men and women. Furthermore, this agrees with widely accepted knowledge on UIP [19]. The rise in UIP likelihood as age increased was also expected based on prior studies [20]. The lack of significance observed in smokers vs. non smokers in the pathological UIP group was unexpected. Significance was observed in the prevalence of focal UIP in smokers vs. non smokers, which was predictable as the criteria for focal UIP positivity was lower than that for pathological UIP; we thus assumed greater stratification between diseased and non-diseased patients. Although a patient cohort comprised entirely of typical UIP positive patients would be expected to display a downward trend in FVC, DLco, and higher KL-6 values in the diseased population, our cohort is made up of many different etiologies, including subacute conditions, and thus the reverse trend in FVC and the lack of significance in DLco and KL-6 was not ultimately surprising [21,22]. The lack of significance in these variables highlights the many confounding factors present in cases of PPF. However, while KL-6 rates between UIP+ and UIP-were not significant, we found that KL-6 was a significant prognostic factor in PPF.

Regarding our histologic criteria, we decided against requiring fibroblastic foci to classify an area as UIP. The focal UIP+ label was defined as destructive fibrosis that did not reach the threshold of NSIP, with a cutoff value of 10% of the diseased area consisting of UIP pattern, as validated by ROC analysis. We also did not require temporal heterogeneity. Prior diagnostic guidelines have stated that it is acceptable to apply the label of UIP even when some of its core histological characteristics are missing, especially when there is a lack of features suggestive of an alternative diagnosis [10,11]. The reason we termed this histology “focal UIP” rather than simply destructive fibrosis was due to prior research mentioning the coexistence of UIP and other ILDs such as CTD-ILD or NSIP - and showing the prognostic significance of such coexistence [7,23,24].

A key finding of this study is that regardless of diagnosed etiology, the presence or absence of UIP had a significant prognostic effect in our patient cohort. Three out of our four disease classifications reached significance in their Kaplan-Meier survival curves. Regarding the one that did not, cHP, it should be noted that the focal UIP-group had a mortality rate of zero and all deaths occurred in the focal UIP+ group. Given the low number of overall cases of cHP in our cohort, statistical tests of this group could be prone to error.

Our finding that different underlying ILD etiologies did not change the overall negative prognostic effect of UIP could be interpreted as support for the “UIP bucket” hypothesis [9]. This hypothesis suggests that all ILDs exhibiting UIP make up the same disease-one which eventually progresses to end-stage honeycomb lung as a phenotype regardless of the initial ILD diagnosis. Instead, varying ILD diagnoses such as HP and CTD-ILD represent accelerating factors of disease progression rather than primacy causes of UIP, and depending on these factors, the distribution of UIP fibrosis varies. This could explain why UIP pattern exhibits worsened prognosis in PPF regardless of the diagnosed etiology. In fact, a prior study has reported that 34% of fibrosing ILDs other than IPF eventually show a progressive phenotype with a prognosis similar to that of IPF [25]. This study also found that UIP was a significant prognostic factor in their 509-patient non-IPF fibrosing ILD cohort.

In terms of clinical suggestions derived from our findings, it is crucial to accurately diagnose PPF cases in which therapeutic agents can have a positive impact. Antifibrotic agents have been proven to be effective in treating patients with PF-ILD [2,26,27]. Since PPF is diagnosed through follow-up within a period of one year, early identification of pathological or focal UIP pattern is valuable for the prompt initiation of treatment, particularly where SLB is widely implemented [28]. A prospective clinical trial is warranted in order to validate these suggestions. Furthermore, for very early stage ILDs, when CT is unclear about the presence of UIP, SLB may be an appropriate recommendation to accurately diagnose pathological and focal UIP. For pathologists without access to digital pathology workflows, judgements based on microscopic evaluation of focal UIP presence must be made. We recommend that if a pathologist feels that UIP pattern may exceed 10% of the disease area, they should advise the clinician to pursue intense follow-up.

This study has multiple limitations. Firstly, it is important to note that this study was conducted at a single institution, and further confirmation from multiple facilities is required to validate these findings. As this study uses SLB as the primacy method of biopsy, our results are most helpful to other institutions which use SLB routinely, but the 2022 guidelines no longer require SLB, so we expect the prevalence of this methodology to decrease [1]. Additionally, due to the subjective nature of pathologists’ interpretations of the UIP pattern and the potential for other histopathological features to serve as prognostic factors, the use of more objective tools, such as image analysis empowered by artificial intelligence may be beneficial in determining these factors more objectively and potentially more accurately.

On the other hand, the strengths of our study lie in that this is the first paper to describe the histology of PPF and relate it to a prognostically significant finding. We also enlisted the expertise of three pulmonary pathologists who blindly evaluated focal UIP, showing high agreement and reproducibility.

The incidence of focal UIP-like fibrosis with a threshold value of 10% has been deemed a crucial criterion for prognosis in PPF; focal UIP like fibrosis that is greater than or equal to 10% tends to indicate a worse prognosis even when controlling for clinical and etiological factors. These findings highlight the significance of identifying and monitoring the presence of focal UIP-like fibrosis even in non-UIP cases. Three expert pathologists have shown a clear correlation between the presence of focal UIP like fibrosis and a less favorable outcome. The focal UIP pattern as defined in this paper is particularly significant, as its rapid recognition can facilitate the prompt initiation of treatment and improve patient outcomes [29,30].

## CONCLUSION

Approximately half of PPF cases exhibit pathological UIP, which correlated with worse survival in our 201-case PPF cohort. Focal UIP, which is defined as UIP pattern occupying 10% or more of the total disease area, has a much higher frequency (∼70%), and its presence is also correlated with a less favorable prognosis. The prognostic significance of focal UIP in PPF holds true regardless of underlying ILD etiology. Even in cases that were negative for pathological UIP but positive for focal UIP, overall survival was significantly worse in the focal UIP+ group.

In conclusion, the histological pattern of UIP in cases of PPF is prognostically meaningful. We hope that this study will spur subsequent research to validate and further explore the connection between the UIP pattern and PPF, and that the histology of PPF may get more recognition in future updates of PPF guidelines. Our study highlights the importance of considering the presence of focal UIP in the evaluation of PPF.

## Author Contributions

Y.T. and E.N.O contributed equally as first authors. Guarantors of integrity of entire study, Y.T., J.F.; study concepts/study design and data acquisition: Y.T., E.N.O, J.F.; analysis/interpretation, all authors; manuscript drafting or manuscript revision for important intellectual content, all authors; approval of final version of submitted manuscript, all authors.

## Data Availability Statement

The data that support the findings of this study are available from the corresponding author upon reasonable request. Code for data cleaning, visualization, and analysis is provided. It is available at https://github.com/eokoshi/PPF.git for review. It can be uploaded to a journal repository upon request once the paper has been conditionally accepted.

## Conflicts of Interest and Source of Funding

All authors declare that they have no conflicts of interest. Funding provided by the Study Group on Diffuse Lung Disease, Scientific Research/Research on Intractable Diseases in the Ministry of Health, Labour and Welfare, Japan.

## Human Ethics Approval Declaration

This study was carried out at Nagasaki University in compliance with the principles of the Declaration of Helsinki and approved by its institutional review board (IRB No. 14012746, February 3, 2014). Patient approval or informed consent was waived because the study involved a retrospective review of patient records.

## SUPPLEMENTAL MATERIALS

**Supplementary Figure 1.**
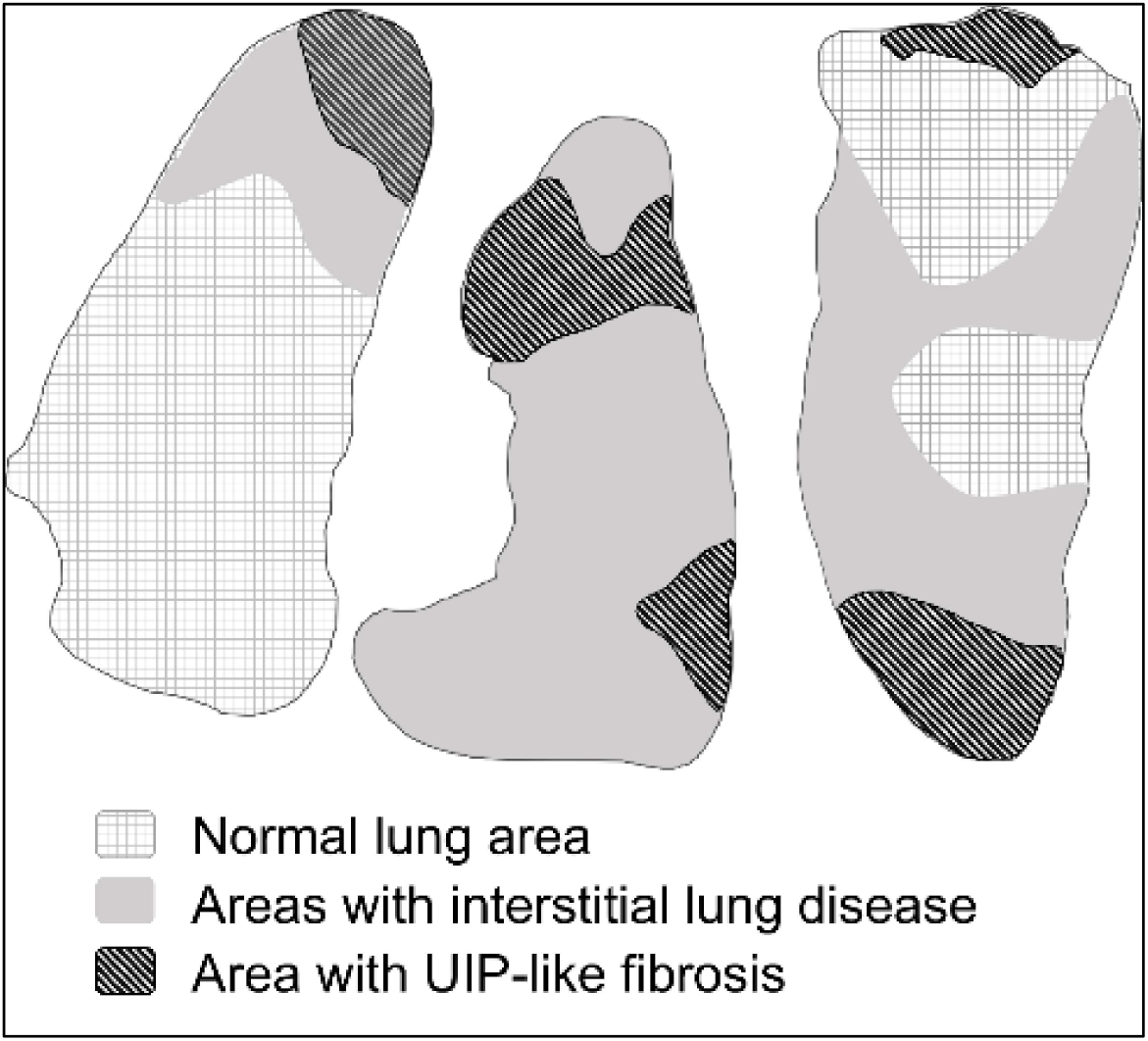
Illustration of focal UIP identification methodology. Representative diagrams of surgical lung biopsy specimen. The areas of UIP-like fibrosis were demarcated using a digital pen tool, and the presence of UIP-like fibrosis within the identified interstitial lung disease areas was quantified separately. The area of UIP-like fibrosis (U) and the total area of ILD (L) were measured, excluding normal tissue area in all specimens obtained through surgical lung biopsy. The focal area of UIP-like fibrosis was defined as U/L, and for cases with multiple biopsy specimens, we took the average percentage across specimens. Focal UIP was defined as UIP covering 10% of the total lesion area. UIP, usual interstitial pneumonia.

**Supplementary Figure 2.**
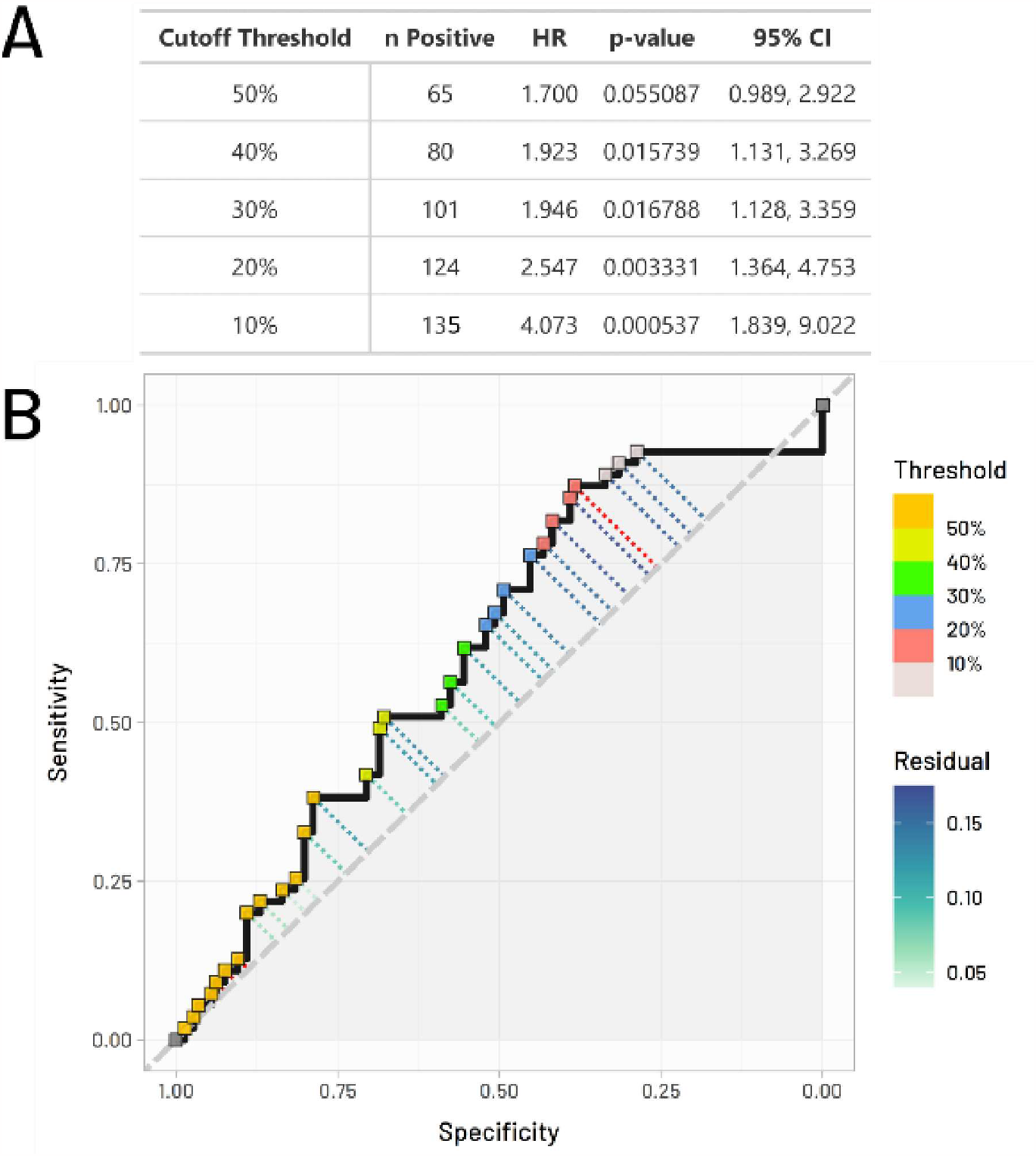
ROC Curve for evaluating cutoff thresholds for focal UIP diagnosis. A) Table showing results of individual fits of a univariate Cox proportional hazards model to each cutoff threshold. A 10% area of UIP within the wider diseased area showed the highest separation between prognostic states. “n Positive” indicates the number of cases positive for focal UIP using that cutoff value. B) ROC curve showing predictive ability of UIP area percentage for death (n = 50) or lung transplant (n = 5) occurrence. Data point with the highest distance from the center line is highlighted with a red residual line (sensitivity = 0.873, specificity = 0.38, threshold = 10.05%). HR, hazard ratio.

**Supplementary Table 1.**
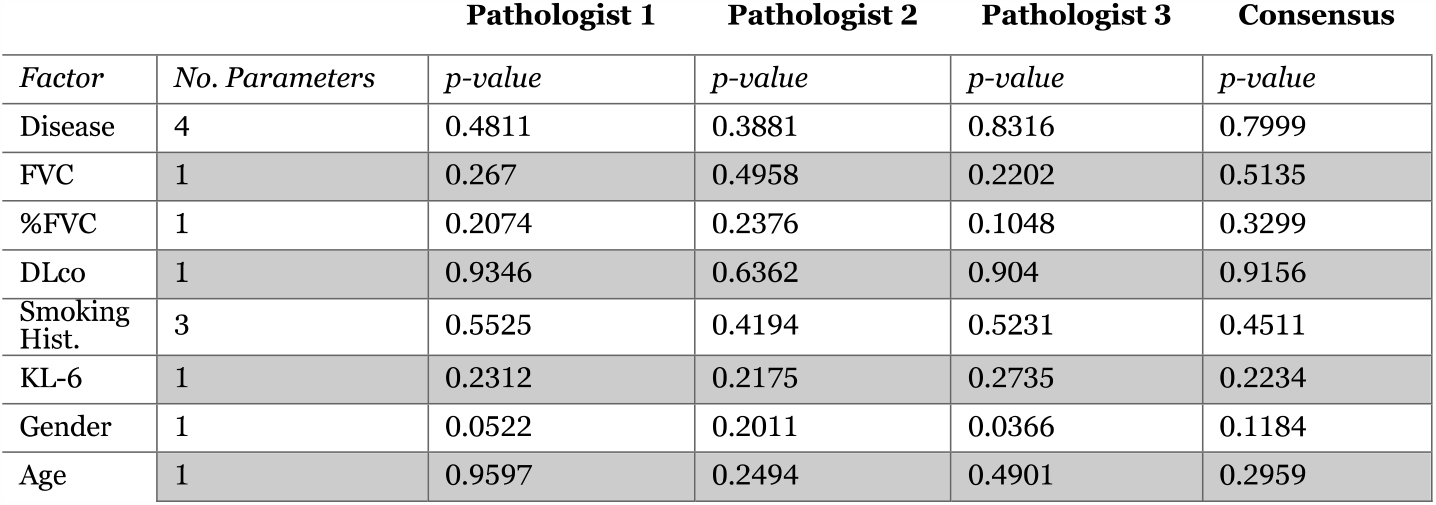
Logistic analysis with individual and consensus results of baseline cohort characteristics’ correlation with survival. Significance tested with likelihood ratio test for effects in logistic analysis. Significance evaluated at p < 0.05. UIP, usual interstitial pneumonia; FVC, forced vital capacity; DLco, carbon monoxide diffusing capacity.

## Notes

### Competing Interest Statement

The authors have declared no competing interest.

### Funding Statement

This study was funded by the Study Group
on Diffuse Lung Disease, Scientific
Research/Research on Intractable Diseases in the
Ministry of Health, Labour and Welfare, Japan.

### Author Declarations

IRB of Nagasaki University gave ethical approval for this work (IRB No. 14012746, February 3, 2014).

### Summary of Updates

Changed the title for accession reasons and deleted a blank page in the pdf.

